# Usability of a Machine-Learning Clinical Order Recommender System Interface for Clinical Decision Support and Physician Workflow

**DOI:** 10.1101/2020.02.24.20025890

**Authors:** Andre Kumar, Jonathan Chiang, Jason Hom, Lisa Shieh, Rachael Aikens, Michael Baiocchi, David Morales, Divya Saini, Mark Musen, Russ Altman, Mary K Goldstein, Steven Asch, Jonathan H Chen

## Abstract

**Objective:** To determine whether clinicians will use machine learned clinical order recommender systems for electronic order entry for simulated inpatient cases, and whether such recommendations impact the clinical appropriateness of the orders being placed.

**Materials and Methods:** 43 physicians used a clinical order entry interface for five simulated medical cases, with each physician-case randomized whether to have access to a previously-developed clinical order recommendation system. A panel of clinicians determined whether orders placed were clinically appropriate. The primary outcome was the difference in clinical appropriateness scores of orders for cases randomized to the recommender system. Secondary outcomes included usage metrics and physician opinions.

**Results:** Clinical appropriateness scores for orders were comparable for cases randomized to the recommender system (mean difference -0.1 order per score, 95% CI:[-0.4, 0.2]). Physicians using the recommender placed more orders (mean 17.3 vs. 15.7 orders; incidence ratio 1.09, 95% CI:[1.01-1.17]). Case times were comparable with the recommender system. Order suggestions generated from the recommender system were more likely to match physician needs than standard manual search options. Approximately 95% of participants agreed the system would be useful for their workflows.

**Discussion:** Machine-learned clinical order options can meet physician needs better than standard manual search systems. This may increase the number of clinical orders placed per case, while still resulting in similar overall clinically appropriate choices.

**Conclusions:** Clinicians can use and accept machine learned clinical order recommendations integrated into an electronic order entry interface. The clinical appropriateness of orders entered was comparable even when supported by automated recommendations.

## Introduction

Physician compliance with evidence-based care often falls short, with overall compliance with clinical guideline recommendations ranging from 20 to 80%.^1^ Such variability may compromise care quality, cost effectiveness, or expedient healthcare delivery, especially when knowledge is inconsistently applied.^2^ The advent of the meaningful use era of electronic health records (EHRs)^3^ creates the opportunity for data-driven clinical decision support (CDS) that utilizes the collective expertise of many practitioners in a learning health system.^4–8^ It may additionally facilitate the acquisition of medical knowledge by enabling clinicians to adopt evolving evidence-based practice patterns.(Holroyd et al. 2007) Tools such as order sets already reinforce consistency and compliance with best practices,^9,10^ but maintainability is limited in scale by a top-down, knowledge-based approach requiring the manual effort of human experts.^11^ Moreover, the intended vs. actual usage of EHR order sets may not align with physician workflows,^12^ and it may impede physicians from learning appropriate alternatives toward patient care.(Kumar and Allaudeen 2016) A key challenge to fulfill a future vision for clinical decision support^13,14^ is the automatic production of content from the bottom-up by data-mining clinical data sources.^15^

Most prior studies in automated development of clinical decision support content have been strictly offline analytical evaluations^5,16,21–25^, with few studies assessing the response of human clinicians to such recommender tools and their ordering patterns. More broadly, the majority of physicians have significant distrust or negative attitudes toward the EHR,^26–28^ which may affect how well these tools could be adopted. As with many machine learning models designed to support clinical decision making, it is unknown if physicians will actually accept such suggestions into their clinical decision workflow.

Previously, we developed a clinical order recommender system by automatically data-mining hospital EHR data.^16^ The results of this approach align with established standards of care^15,17,18^ and is predictive of real physician behavior and patient outcomes.^16^ Our underlying vision is to seamlessly integrate a system into clinical order entry workflows that automatically infers the relevant clinical context based on data already in the EHR and provides actionable decision support in the form of clinical order suggestions, analogous to Netflix or Amazon.com’s “customers who bought A also bought B” system.^19,20^ It is unknown if these suggested orders would improve the quality of care and be readily accepted into clinical decision making.

This study seeks to address these issues by examining physicians’ behaviors while interacting with a clinical provider order entry (CPOE) interface that simulates an electronic health record for hospital clinical scenarios. We specifically examine whether the clinical recommender system impacted the number of clinically inappropriate/appropriate orders placed during the simulated encounters. We also add expanded results related to physician ordering patterns, user experience metrics, and survey responses when a clinical order recommender system is added to standard functionality.

### Objective

To determine whether clinicians will use machine learned clinical order recommender systems for electronic order entry for simulated inpatient cases, and whether such recommendations impact the clinical appropriateness of the orders being placed and the system’s impact on physician workflow.

## Methods

### Participants and Setting

This study was conducted at a single academic institution from 10/2018-12/2019. We recruited physicians (n=43) with experience caring for medical inpatients within the past year using local mailing listservs. Participants included both medical residents (trainees who have a medical license but still require oversight) and supervising physicians. The study was approved by the Stanford University Institutional Review Board.

### Study Design & Outcomes

Participants were offered a $195 incentive payment for a 1 hour usability testing session in a closed office setting where they were exposed to a series of five clinical cases that simulate common inpatient medical problems (see *Cases & Grading* below) on a digital interface that simulated their institution’s electronic health record. Upon recruitment to the study, a researcher guided participants through two demonstration cases (diabetic ketoacidosis and chest pain) to illustrate basic functions of the digital interface (data review, order entry, order sets). The subsequent five cases were presented to participants in a sequence randomly assigned for each user (Table 2).

All participants were randomized to undergo each of the five cases with either an available clinical recommender system that offered order suggestions vs. no recommender system. Conventional clinical order entry options including order set checklists and manual search of individual orders by name were available in all cases, making usage of the recommender system completely optional compared to their usual order-entry workflows. Participant activity was recorded through screen capture, audio, and user interface tracking software. Following the case series, all participants filled out a survey on their experiences with the system and their receptiveness to a clinical recommender system.

Outcome measures included the time to complete the case, the number of clinical orders selected from manual search vs. the automated recommender system, usage metrics (e.g. number of clicks), clinical quality of orders placed (see *Cases & Grading*), and survey results.

### Clinical Recommender Development

As described previously,^16^ we extracted deidentified structured data for all inpatient hospitalizations from the 2009-2014 STRIDE clinical data warehouse.^29^ The data cover >74K patients with >11M instances of >27K items (medication, laboratory, imaging, and nursing orders, lab results and diagnosis codes). We built a clinical collaborative filtering (recommender) system based on this data, modeled on Amazon’s product algorithm^19,20^ using item co-occurrence statistics.

We built a simulated computerized physician order entry (CPOE) interface with open technologies including PostgreSQL, Python, Apache HTTP, and HTML/JavaScript. Our unique addition is an automated recommender (Figure 1), analogous to a “Customers Who Bought This Item Also Bought This…” service that anticipates other clinical orders that are likely to be relevant based on similar prior cases in prior electronic health records.

**Figure 1.**
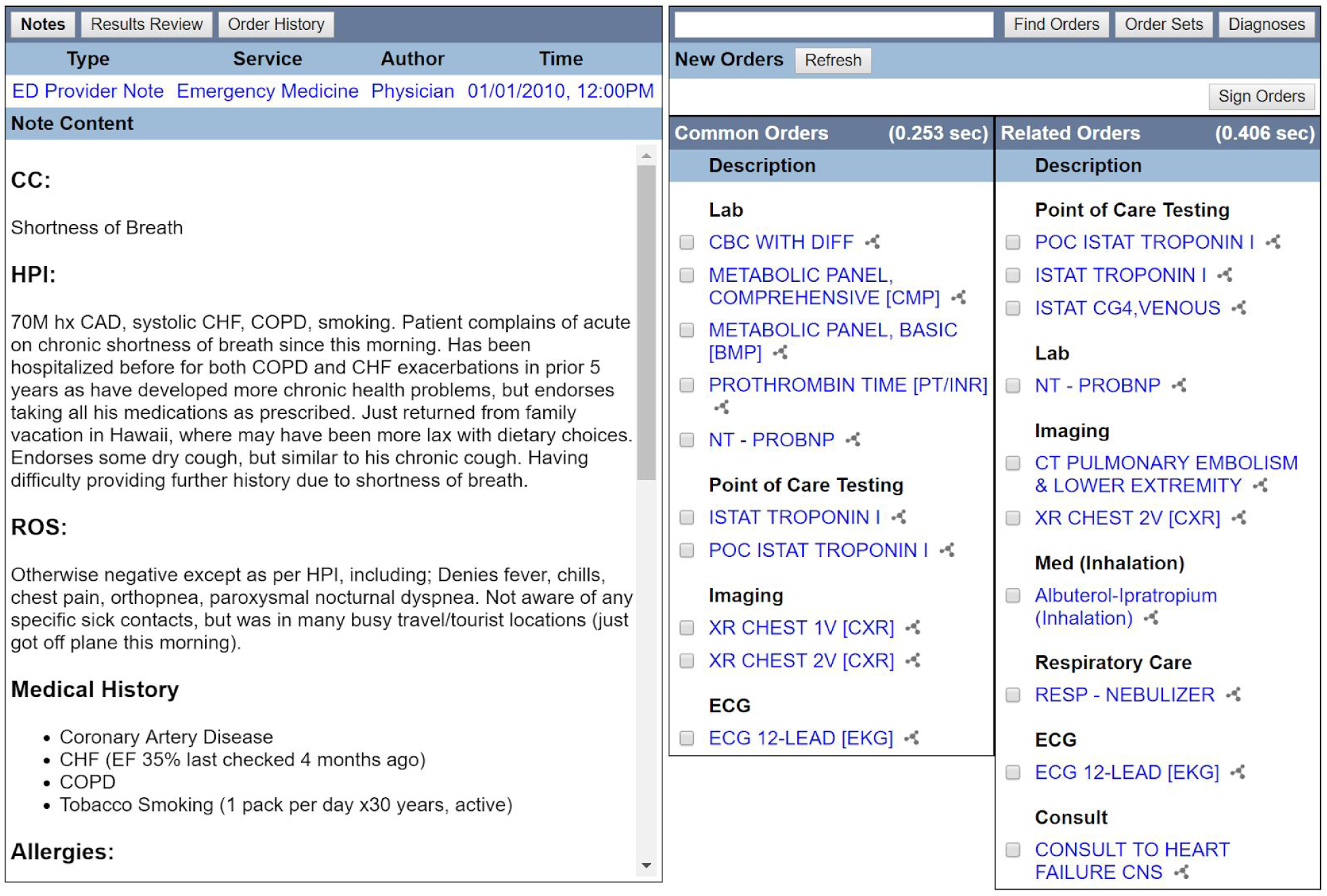
Simulated clinical order entry interface, notes and clinical order recommender. Standard functions include navigation links to review notes and results (top-left). Order entry includes a conventional search box for individual orders and pre-authored order sets (top-right). A recommender algorithm suggests clinical orders (right), in this example triggered by a presenting symptom code (Shortness of Breath, ICD9 786.05). Clinical orders predicted most likely to occur next are highlighted under *Common Orders*, while those under *Related Orders* are less likely but disproportionately associated with similar cases and thus may be more specifically relevant. As users enter additional orders, the recommender algorithm continually updates the suggested lists based on the accumulating information.

**Figure 2.**
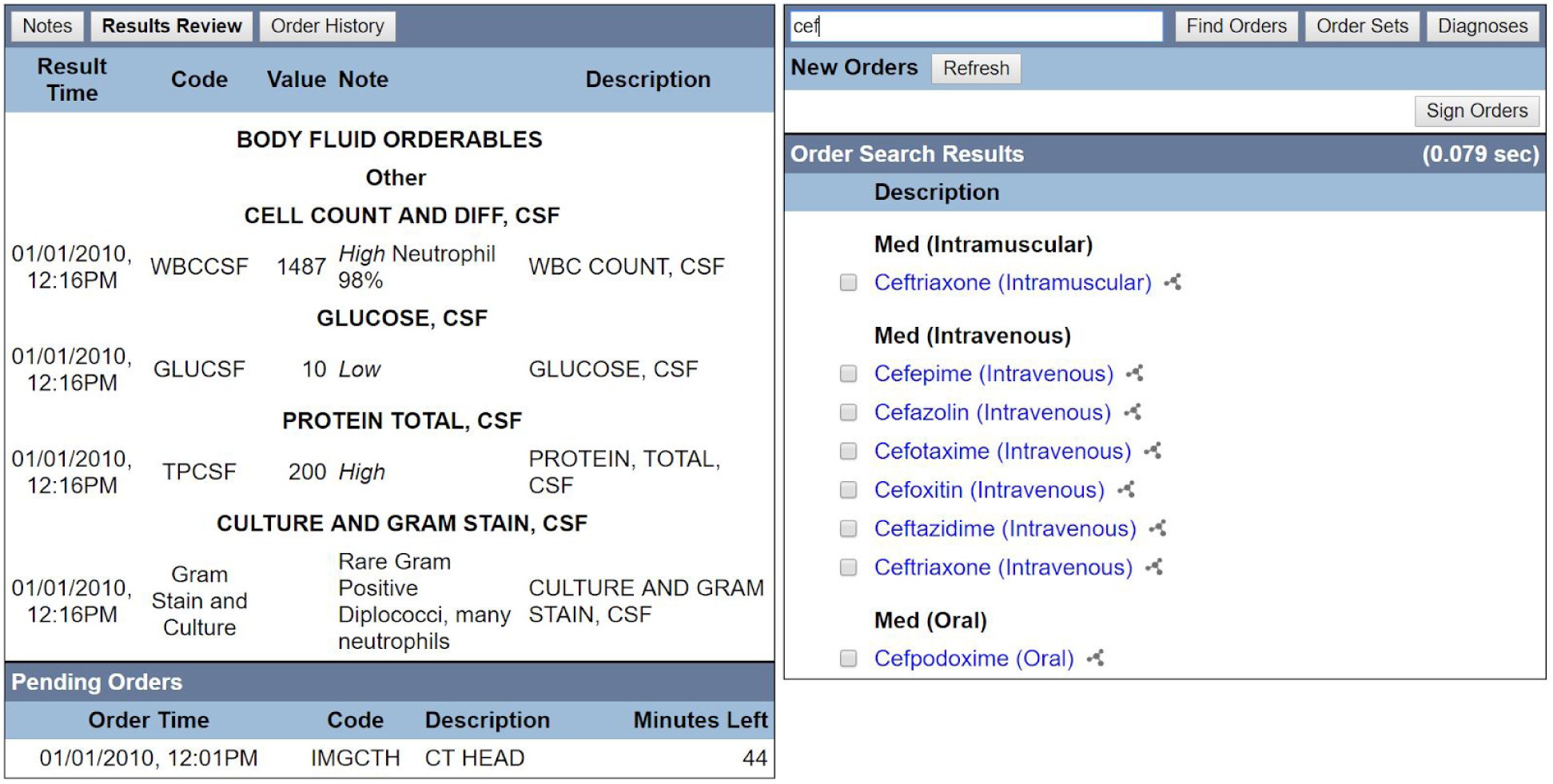
Simulated clinical order entry interface, results review and clinical order manual search. Users can order diagnostics such as cerebrospinal fluid (CSF) studies to review results (left). This may require simulated passage of time for results to be ready (e.g., CT Head requiring another 44 minutes for results to be ready). Conventional manual search for clinical orders via a text search box (top-right) yields clinical order options (right) identified by prefix. In this example, identifying all clinical orders with a word starting with “cef.”

### Cases & Grading

A panel of board-certified internal medicine physicians (AK, JH, LS, and JHC) developed 5 clinical cases of common inpatient medical problems: unstable atrial fibrillation, neutropenic fever, variceal gastrointestinal hemorrhage, bacterial meningitis, and acute pulmonary embolism (Table 1). Each participant was exposed to the clinical interface (Figure 1), which included the patient’s history and physical examination. Depending on the interventions ordered, the case would progress across several decisional nodes (Table 1). For example, if a participant ordered a lumbar puncture and antibiotics for bacterial meningitis, the case would progress toward a different node (patient improvement). In contrast, if antibiotics were not ordered, the patient would deteriorate (updated vitals and clinical notes would appear in this node). Diagnostic test results are only visible and change state if respective orders are entered (e.g., low hemoglobin revealed only if a blood count is ordered and changes if a blood transfusion is ordered). With each order entered, the clinical order recommender lists updates based on the accumulating patient information.

**Table 1.**
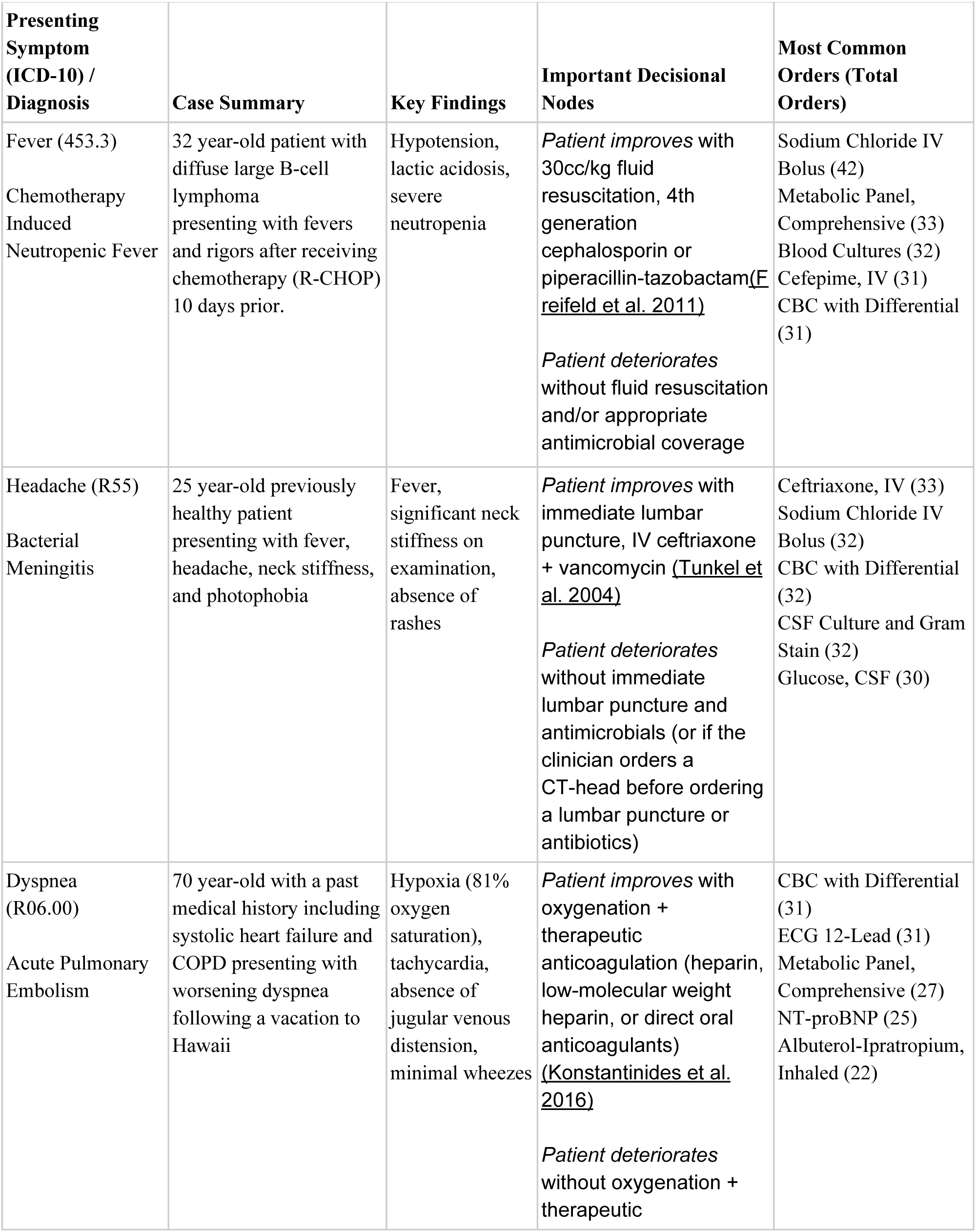
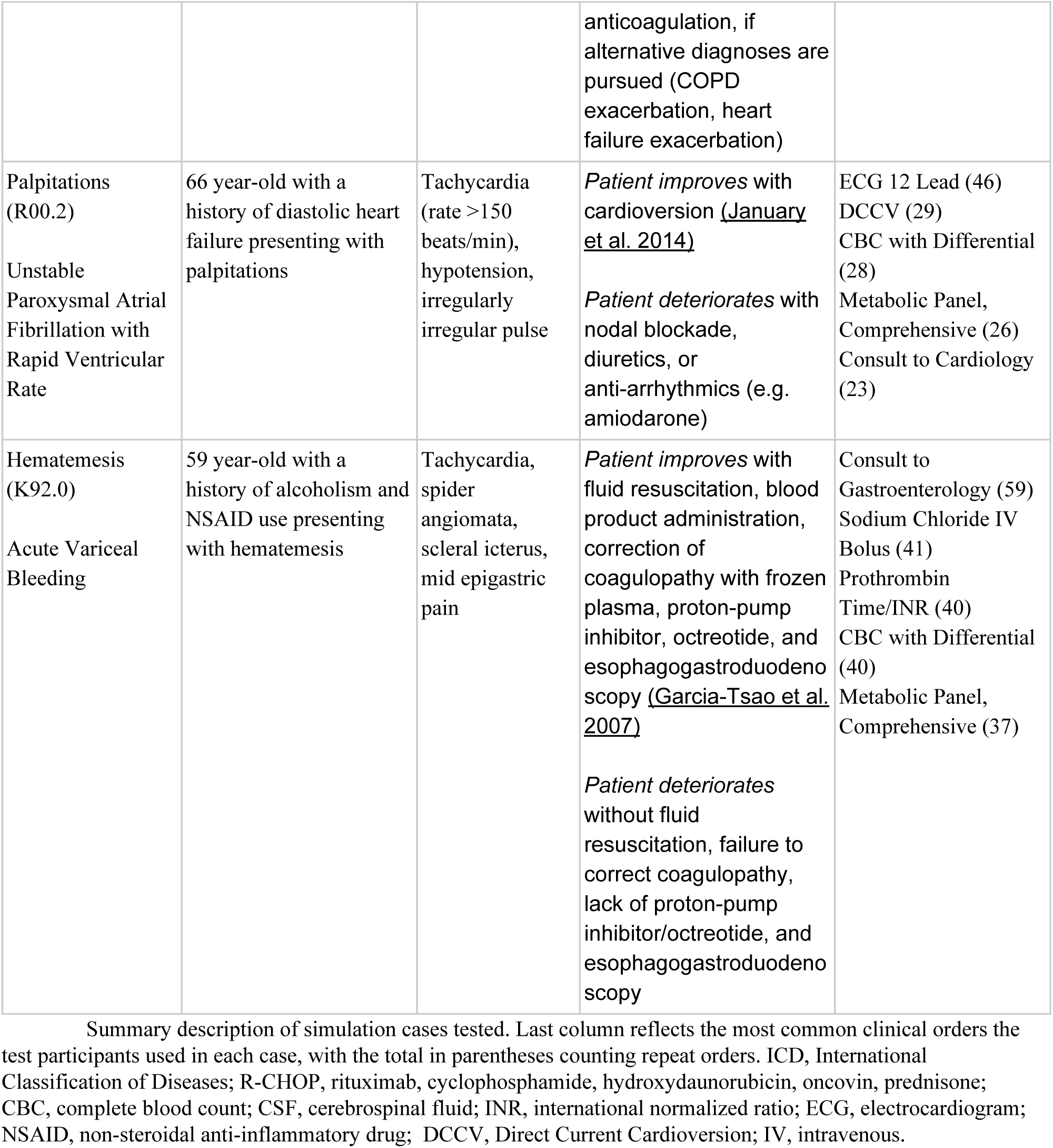
summarizes key elements of the five case scenarios that participants were tested with.

Delphi method was used by the case designers to determine clinical appropriateness for all orders. (Hsu and Sandford 2007) Following the creation of this system, each physician (AK, JH, LS, and JHC) independently reviewed all orders placed for the case. Cases were classified according to their state (initial, subsequent, or resolution) and orders were considered in the context of each state. Based on prior consensus on the scoring procedure, each grader assigned an individual score to an order on a −10 (very harmful) to +10 (very beneficial) scale. Please see the Appendix for a full description of how grading was considered. The reviewers achieved an initial intraclass correlation coefficient (ICC) of 0.51 (95% CI: [0.47-0.53]) for all scored orders on a −10 to +10 scale (https://www.ncbi.nlm.nih.gov/pmc/articles/PMC4913118/). Following independent scoring of each order, the reviewers met as a group to review their scores. Appropriate research studies and clinical guidelines were considered when assigning a consensus score (Table 1). For items that did not have perfect interrater agreement, the group convened and deliberated to assign a consensus score.Hsu and Sandford 2007 In instances where the panel could not reach a final consensus, no score was assigned and the order was not included in the final analysis.

### Case-Based Scenarios

## Results

### Participants

A total of 43 physicians participated in this study, with a total of 215 unique observations. The physicians had a median of 3.0 [IQR: 3.0-5.0] years since obtaining their medical degree. Approximately 30 (70.0%) identified their primary specialty as Internal Medicine, 8 (18.6%) identified Emergency Medicine, 25 (58.1%) were resident trainees, and 19 (44.1%) were board certified in their respective specialty.

### Primary Outcome

The mean assigned score per order for all participants was 6.2 (95% CI: [4.8, 7.5]; Table 1). There was no significant difference detected in the mean score per order for physicians randomized to the clinical recommender (mean 0.1 decrease in score, 95% CI: [-0.4, 0.2]). Random effects modeling for physicians revealed a SD of 0.4 (95% CI: [0.1-0.6]), while random effects modeling for the clinical cases revealed an SD of 1.4 (95% CI: [0.7-2.7]), suggesting most variations in scores occurred due to the clinical cases rather than the participants.

### Secondary Outcomes

#### A. Physician Experience

Overall, physicians spent an average time of 6.9 (standard error, 0.6) minutes per clinical module, with a mean 55.2 (standard error, 3.9) navigation clicks between sections (e.g., notes vs. results review) and 16.7 (standard error, 0.9) clinical orders per case. (Table 3). Physicians randomized to the recommender had approximately 10% less navigational clicks (mean clicks 53.1) compared to physicians without the recommender (mean clicks 58.4; incidence ratio 0.90, 95% CI: [0.83-0.99]). Physicians ordered a greater amount of orders when the recommender system was available (mean 17.3 vs. 15.7 orders per case; incidence ratio 1.09, 95% CI: [1.01-1.17]). Across different simulated case types, there was not a consistent trend in physicians taking more or less time to complete cases with the recommender system available (Table 3). In subgroup analysis of resident physicians in training vs. non-residents (Appendix Material), there appeared to be varying effects with residents spending less case time and ordering more with the recommender available, while non-residents spent more case time.

**Table 3a.** Usage metrics when clinical order recommender system was available vs. not. Reported as totals, proportions, or means +/− standard error. Options reflects clinical order options that were presented to the user for consideration via either manual search results or automated recommender. Recommender precision (positive predictive value) reflects the proportion of clinical order options from the recommender that were actually used. Clinical benefit, harm or neutrality was based on the integer assigned by the expert panel consensus (e.g. positive, negative, or zero) for each order in the context of each clinical case.

| All Cases |  | P |
| --- | --- | --- |
| No | Yes | Recommender Available |
| 86 | 129 | Number of Cases |
| 6.7+/-0.6 | 7.1+/-0.6 | Case Time (Minutes) |
| 58.4+/-4.1 | 53.1+/-3.7 | Navigation Clicks |
| 15.6+/-0.9 | 17.3+/-0.8 | Clinical Orders, Total |
| 15.6+/-0.9 | 7.3+/-0.7 | Orders from Manual Search |
| 81.5+/-8.1 | 44.5+/-5.5 | Options from Manual Search |
| N/A | 10.0+/-0.6 | Orders from Recommender |
| N/A | 40.0+/-1.0 | Options from Recommender |
| 19% | 16% | Manual Search Precision |
| N/A | 25% | Recommender Precision |
| N/A | 42% | Manual Search Recall |
| N/A | 58% | Recommender Recall |
| 84.3+/-4.3 | 89.8+/-4.4 | Total Score |
| 6.4+/-0.2 | 6.0+/-0.3 | Score per Order |
| 11.1+/-0.5 | 12.2+/-0.5 | Clinically Beneficial Orders per Case |
| 0.3+/-0.1 | 0.6+/-0.1 | Clinically Harmful Orders per Case |
| 2.4+/-0.4 | 2.4+/-0.3 | Clinically Neutral Orders per Case |

**Table 3b.** Usage metrics when clinical order recommender system was available vs. not, separated by Resident physicians (trainees) vs. non-Residents. Reported as totals, proportions, or means +/− standard error. Clinical benefit, harm or neutrality was based on the integer assigned by the expert panel consensus (e.g. positive, negative, or zero) for each order in the context of each clinical case.

| Non-Residents |  | Residents (Trainees) |  |  |
| --- | --- | --- | --- | --- |
| No | Yes | No | Yes | Recommender Available |
| 38 | 57 | 48 | 72 | Cases |
| 7.1+/-0.5 | 8.1+/-0.7 | 6.4+/-0.8 | 6.3+/-0.5 | Case Time (Minutes) |
| 60.6+/-4.8 | 55.3+/-4.3 | 56.6+/-4.4 | 51.3+/-4.0 | Navigation Clicks |
| 16.7+/-1.1 | 17.4+/-1.0 | 14.8+/-0.9 | 17.3+/-0.7 | Clinical Orders, Total |
| 16.7+/-1.1 | 7.5+/-0.8 | 14.7+/-0.9 | 7.2+/-0.8 | Orders from Manual Search |
| 81.0+/-9.0 | 43.2+/-5.8 | 81.9+/-9.2 | 45.5+/-6.4 | Options from Manual Search |
|  | 9.8+/-0.7 |  | 10.2+/-0.7 | Orders from Recommender |
|  | 39.7+/-0.9 |  | 40.3+/-1.3 | Options from Recommender |
| 21% | 17% | 18% | 16% | Manual Search Precision |
|  | 25% |  | 25% | Recommender Precision |
|  | 43% |  | 42% | Manual Search Recall |
|  | 57% |  | 59% | Recommender Recall |
| 86.4+/-6.1 | 89.8+/-7.2 | 82.7+/-5.9 | 89.8+/-5.4 | Total Score |
| 6.2+/-0.4 | 6.0+/-0.4 | 6.5+/-0.3 | 6.1+/-0.4 | Score per Order |
| 11.6+/-0.8 | 12.2+/-0.8 | 10.8+/-0.6 | 12.2+/-0.5 | Clinically Beneficial Orders per Case |
| 0.4+/-0.2 | 0.6+/-0.2 | 0.3+/-0.1 | 0.6+/-0.2 | Clinically Harmful Orders per Case |
| 2.8+/-0.7 | 2.4+/-0.4 | 2.0+/-0.4 | 2.4+/-0.2 | Clinically Neutral Orders per Case |

All physicians used clinical order options from the recommender system at least once, including 127 (98%) of the 129 physician-cases where the recommender was available. This corresponded to much less need for manual searching for clinical orders with the number of available order options considered from manual searches being 54% less when the recommender system was available vs. unavailable (mean 44.5 vs. 81.5). The *recall* of the recommender options was consistently greater than manual search options (58% vs 42%), indicating users were more likely to find the clinical orders they wanted from the automated recommender lists than from options returned by manual search. The *precision* of the recommender options was similarly greater than manual search options (25% vs 16%), indicating users had to sift through fewer irrelevant options to find the clinical orders they wanted than the number of irrelevant options produced by manual searches.^12^

#### B. Scoring Outcomes

Clinician-cases randomized to the clinical recommender had a 6% increase in the total scores per case (incidence ratio 1.06 (95% CI: [1.01-1.12]). The number of clinically beneficial orders, defined as a positive integer on the grading scale for a given case, demonstrated a trend toward statistical significance when the recommender was available vs. unavailable (mean number of positively-graded orders 12.2 vs. 11.1; incidence ratio 1.080, 95% CI: [0.99-1.17]). There was no difference in the number of clinically neutral or harmful orders for when the recommender was available vs. unavailable (mean number of negative/neutral orders 3.0 vs. 2.7; incidence ratio 0.99, 95% CI: [0.81-1.22])..

**Appendix Table A.** Usage metrics stratified per simulated case scenario for when the clinical order recommender system was available vs. not. Reported as totals, proportions, or means +/− standard error. Clinical harm, benefit, or irrelevance was based on the integer assigned by the expert panel consensus for each order in the context of each clinical case.

| Atrial Fibrillation, Unstable |  | Gastro-Intestinal Bleed |  | Meningitis, Bacterial |  | Neutropenic Fever |  | Pulmonary Embolism |  | Case Description |
| --- | --- | --- | --- | --- | --- | --- | --- | --- | --- | --- |
| No | Yes | No | Yes | No | Yes | No | Yes | No | Yes | Recommender Available |
| 17 | 26 | 17 | 26 | 12 | 31 | 22 | 21 | 18 | 25 | Number of Cases |
| 4.6 | 6.6 | 9.1 | 8.5 | 6.7 | 5.4 | 4.4 | 5.3 | 9.3 | 9.7 | Case Time (Minutes) |
| 47.2 | 48.3 | 83.9 | 74.4 | 53.3 | 40.2 | 36.9 | 36.0 | 74.4 | 66.2 | Navigation Clicks |
| 11.5 | 14.6 | 21.5 | 22.4 | 19.2 | 16.3 | 12.1 | 13.8 | 16.1 | 19.2 | Clinical Orders, Total |
| 11.5 | 6.0 | 21.6 | 9.4 | 18.8 | 7.8 | 12.1 | 3.0 | 16.1 | 9.5 | Orders from Manual Search |
| 53.5 | 27.1 | 127.8 | 59.8 | 70.0 | 41.1 | 56.6 | 33.4 | 102.3 | 60.0 | Options from Manual Search |
|  | 8.7 |  | 12.8 |  | 8.4 |  | 10.7 |  | 9.9 | Orders from Recommender |
|  | 36.5 |  | 45.6 |  | 38.5 |  | 35.6 |  | 43.6 | Options from Recommender |
| 21% | 22% | 17% | 16% | 27% | 19% | 21% | 9% | 16% | 16% | Manual Search Precision |
|  | 24% |  | 28% |  | 22% |  | 30% |  | 23% | Recommender Precision |
|  | 41% |  | 42% |  | 48% |  | 22% |  | 49% | Manual Search Recall |
|  | 59% |  | 57% |  | 52% |  | 78% |  | 52% | Recommender Recall |
| 49.6 | 49.6 | 121.3 | 120.4 | 85.8 | 86.8 | 74.1 | 88.8 | 93.8 | 104.2 | Total Score |
| 8.8 | 9.3 | 14.1 | 14.5 | 18.4 | 15.6 | 9.1 | 11.0 | 11.9 | 13.9 | Clinically Beneficial Orders per Case |
| 0.9 | 1.6 | 0.2 | 0.3 | 0.7 | 0.7 | 0.0 | 0.0 | 0.0 | 0.0 | Clinically Harmful Orders per Case |
| 1.4 | 2.7 | 2.7 | 2.3 | 5.0 | 2.9 | 1.3 | 0.6 | 2.5 | 3.3 | Clinically Irrelevant Orders per Case |

### Survey Responses

Overall, the clinical decision tool was positively received by the study participants, where 96% agreed or strongly agreed that the tool would be useful for their position. Moreover, 90% agreed or strongly agreed that the system would make their job easier and 86% felt that it would increase their productivity. Thematic analysis revealed a dichotomy in how physicians viewed the system could be used. Approximately 58% of the physicians self-identified the system would be useful for patients who have a clear diagnosis or whose clinical problems could be guided by a step-wise, algorithmic approach. However, a sizable minority (23%) stated the system would be more useful for patients presenting to the emergency department without a clear diagnosis, as this would facilitate expedient ordering of several diagnostic tests to help differentiate the patient. Others mentioned the tool’s utility for diseases that may require several simultaneous orders (for example, diabetic ketoacidosis). Additional comments indicate physicians felt that the tool would be less useful for sub-specialized care or for patients that require few simultaneous orders.

**Table 4.** Physician Survey Responses. Responses were assessed based on a 5-point Likert scale (1 = Strongly Disagree, 2 = Disagree, 3 = Neutral, 4 = Agree, 5 = Strongly Agree)

| Survey Question | 1 | 2 | 3 | 4 | 5 |
| --- | --- | --- | --- | --- | --- |
| I would find the system useful in my job | 0% | 0% | 5% | 49% | 47% |
| Using the system would make it easier to do my job | 0% | 5% | 5% | 44% | 46% |
| This system would increase my productivity | 0% | 9% | 5% | 42% | 44% |
| This system would let me complete tasks more quickly | 0% | 5% | 7% | 37% | 51% |
| This system would increase my job performance | 0% | 7% | 14% | 47% | 32% |

## Discussion

In this study, we found that the use of a clinical order recommender system for common clinical scenarios seen in hospital medicine and emergency medicine did not adversely affect patient care by suggesting more clinically adverse or irrelevant orders. The recommender system did not affect the amount of time physicians spent on an EHR interface, but did it reduce the number of clicks per case. Although physicians placed more orders with the recommender system, this effect was minimal (mean 1.6 more orders per encounter). Importantly, physicians placed less orders from manual searches as a result of the tool. The recommender demonstrated superior recall of orders, suggesting that users were more likely to find the orders they wanted from the recommender rather than from manual searches. The tool was positively received by the study participants, who identified clear benefits toward their workflow and productivity. This represents a key study to examine the use of clinical recommender decision support tools on physician ordering habits and patient care as applied toward inpatient emergency clinical scenarios.

There is wide variability in clinical practice, even in instances where are clear guideline-directed diagnostic and treatment algorithms for well-defined clinical problems.^1^ Such variability may compromise care quality, cost effectiveness, or expedient healthcare delivery.^2^ Healthcare systems have sought to improve both the quality of patient care and the EHR experience by providing standardized order sets.^30–33^ However, order sets may not align with individual cases with many extraneous, irrelevant, or contextual order options.^(Lietal.2019;KumarandAllaudeen2016)^ They are also static: for instance, the manually-authored static order sets available in our own institution for deep venous thrombosis treatment still recommend warfarin therapy, despite direct oral anticoagulants largely becoming the current standard of practice.^42^ Guidance for up-to-date medical care clearly must come from multiple sources, and this points to one potential advantage of the collaborative filtering approach in that it can rapidly and automatically adapt to newly emerging practices.

Our recommender tool essentially functions as a dynamical clinical order set that continuously updates in response to new patient information, demonstrating increased accuracy and reduced need for conventional manual searches. While there are challenges to designing and maintaining such a system, there may be several benefits, including increased physician acceptance and usage (100% of physicians in this study who had the recommender available to them used despite being completely optional). The recommender system interface received largely positive views by our participants, suggesting that physicians will accept machine generated clinical order tools if they are embedded into clinical workflows. More importantly, our expert panel found no significant deterioration in the quality of the clinical orders, and physicians using the recommender system were not more likely to place clinically harmful orders. Notably, there remained substantial variability in the amount of clinically appropriate orders placed by different providers with or without the recommender system. These findings highlight the ongoing variability of clinical practice among physicians (even when additional point-of-care tools are given to them). While order sets have been shown to promote cost-effectiveness,^38,39^ further evaluation is needed to determine how much clinical recommender systems are promoting improved care with more useful orders vs. reducing cost-effectiveness with more unnecessary orders.

Time motion studies indicate that clinicians spend most of their time in the EHR,^34,35^ with many spending significant time searching for and entering orders.^36^ While this study showed a reduction on reliance of manual searches and navigational clicks, interestingly, it did not show a reduction in the amount of time that physicians spent per simulated case. The simulated test setting may have led participants to artificially fill the time within cases, or perhaps the reduction in manual search efforts freed their cognitive attention to attend more to the medical decision making tasks of each case. Additionally, other authors have shown that most of a clinicians time in the EHR is spent in data review (reviewing clinical notes, laboratory results, or diagnostic reports)(Chi et al. 2019; Wang et al. 2019; Ouyang et al. 2016; Ouyang et al. 2016), which was simplified in these clinical scenarios. For example, these patients had only a few notes, compared to many patients who may have hundreds. Future studies should consider the implementation of clinical recommender systems in real practice environments to assess whether they result in time savings for physicians navigating the EHR without compromising quality of care.

There are several limitations to this study. Our tool was based on a clinical data warehouse of electronic health records data that may not be available at all institutions. Similarly, the lack of a broadly accepted open architecture that allows for custom workflow integrations into common commercial EHRs, limits the ease of implementation of the system components studied. Our users were given an orientation of the recommender system and its purpose before engaging with the practice scenarios, which likely contributes a Hawthorne effect on how users interacted and viewed the system. Each testing session was pre-scheduled for a fixed time (1 hour for 5 test cases), which may have artificially constrained the variability in task completion time outcome. Finally, although our expert panel used previously validated methodology to devise a scoring system(Hsu and Sandford 2007), there was still moderate interrater agreement for some clinical orders, which limits the generalizability of these findings to other clinical scenarios or healthcare settings.

At a time when the EHR is met with distrust and negativity by clinicians from the burdens of documentation and data entry, clinical recommender systems represent a key opportunity to improve the quality, consistency, and experience of healthcare. This study represents an important step towards a future where EHRs anticipate clinical needs without even having to ask, so that clinicians can start to feel like the computers are working for them, instead of the other way around.

## Conclusions

Clinical order suggestions from a data-driven recommender system were readily used and accepted by physicians across a variety of simulated inpatient clinical scenarios. This system mildly increased the number of clinical orders placed per case without compromising the number of clinically harmful or clinically irrelevant orders. Physicians were more likely to find the clinical orders they wanted using such tools as compared to manual search methods (i.e. superior precision and recall), but it did not change the overall amount of time they spent in a simulated EHR setting. Nonetheless, clinicians overall view such clinical recommender systems positively, perceiving a clear potential benefit toward their workflow.

## Data Availability

Study data will be shared online.

## lAcknowledgements

This research was supported in part by the NIH Big Data 2 Knowledge initiative via the National Institute of Environmental Health Sciences under Award Number K01ES026837, the Gordon and Betty Moore Foundation through Grant GBMF8040, and a Stanford Human-Centered Artificial Intelligence Seed Grant. This research used data or services provided by STARR, STAnford medicine Research data Repository,” a clinical data warehouse containing live Epic data from Stanford Health Care (SHC), the University Healthcare Alliance (UHA) and Packard Children’s Health Alliance (PCHA) clinics and other auxiliary data from Hospital applications such as radiology PACS. The STARR platform is developed and operated by Stanford Medicine Research IT team and is made possible by Stanford School of Medicine Research Office. The content is solely the responsibility of the authors and does not necessarily represent the official views of the NIH, VA, or Stanford Healthcare.

## Appendix: Determination of Clinical Benefit/Harm of Orders

### Instructions

*The following instructions were given to each of the members of the expert panel to score orders for each clinical case. These instructions were given during the initial scoring phase of the project. Following independent grading by each member of the expert panel, the group convened to assign a consensus score for items that did not have complete interrater reliability. In instances where no consensus could be assigned, a score was not assigned*.

#### Overview of Columns

Each column is separated by the following:

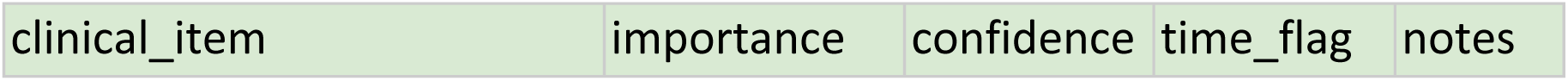

**Clinical_Item:** Which corresponds to a particular order

**Group:** Corresponds to a category of orders (antibiotics, imaging, fluids, medications, diagnostics). Use your best judgement as to how you would group them.

**Importance (Benefit vs Harm):** This corresponds to whether an order is good, bad, or neutral. Graded on a −10 (extremely harmful) to +10 (extremely beneficial) scale to give a gradation.

**Confidence:** How confident an individual is with their score for an item (1-5 Scale). 1= not confident at all; 2-unconfident; 3=neutral; 4=confident; 5= very confident.

**Time_Flag**: Relates to the important decisional nodes for each case.

**Notes:** Make any notes about how you scored this case and why

#### Example

Imagine you had a male patient coming in with nonpurulent cellulitis of the lower extremity with severe sepsis/hypotension. Below are some sample orders:

|  |
| --- |
| Cefazolin 1g IV |
| POC ISTAT, VENOUS BLOOD GASES AND LACTATE (CG4)<br>[VBG] |
| POC URINE HCG QUAL |
| POC URINE DIPSTICK |
| Lisinopril 40mg |

Some orders (Istat blood gases + lactate and IV Cefazolin) might be more relevant/beneficial.

- You would score IV cefazolin as +10 (extremely beneficial), decide how important a lactate would be, and score a −10 for starting a new high-dose blood pressure medication in a patient with an active infection and severe sepsis.
- A pregnancy test is neutral (score of 0) because it is clinically irrelevant.

### Consider harm of procedures

If a patient with well-controlled atrial fibrillation presents to clinic and is currently in sinus rhythm, ordering a DCCV would be harmful without causing immediate benefit (hence the score should be negative).

### Exclude cost considerations

when making a decision (including unnecessary labs)

### Exclude reputational or cultural aspects

It might be ill-advised in real life to consult a pulmonologist for a patient with acute DKA without pulmonary findings, but if it was not immediately relevant to the case, assign 0 points (rather than negative for wasting a hospital resource).

#### Example 2

70 year old patient presenting with cough, fevers, and shortness of breath (e.g. presenting with suspected community acquired pneumonia). The patient gets worse if the provider doesn’t initiate initial antibiotics and fluids.

SAMPLE Orders for the Case

|  |
| --- |
| CEFTRIAXONE 1g IV |
| X-RAY CHEST, 2-VIEW |
| NORMAL SALINE 1 LITER IV |
| METOPROLOL SUCCINATE 10MG IV |
| TRANSESOPHAGEAL ECHO |

| clinical_item | group | importance | confidence | time_flag | Notes |
| --- | --- | --- | --- | --- | --- |
| CEFTRIAXONE 1g IV | antibiotics | +10 | 5 | Initial | +10 points in first part, +5 points in second part for delay (½ points) |
| NORMAL SALINE 1 LITER IV | fluids | +10 | 5 | Deterioration | +10 points in first part, +5 points in second part for delay (½ points) |
| METOPROLOL<br>SUCCINATE 10MG<br>IV | Rate<br>contr<br>olling | -10 | 4 |  | Same throughout |
| TRANSESOPHAGEA<br>L ECHO | imagi<br>ng | -5 | 5 |  | Same throughout |

In order to prevent participants who delayed appropriate care from getting scored the same as other individuals, half points should be awarded for delays in care (e.g. the case progresses to a new decisional node).

Certain orders may have consistent grading across all clinical nodes (they may be clinically irrelevant or always harmful).

